# The rapid reemergence of seasonal respiratory viruses in Houston, Texas, after relaxing COVID-19 restrictions

**DOI:** 10.1101/2021.05.27.21257940

**Authors:** Parsa Hodjat, Paul A. Christensen, Sishir Subedi, David W. Bernard, Randall J. Olsen, S. Wesley Long

**Author notes:** Address correspondence to S. Wesley Long, M.D., Ph.D., Department of Pathology and Genomic Medicine, Houston Methodist Research Institute, 6565 Fannin Street, Suite B490, Houston, Texas 77030. Disclosures: None.

## Abstract

Implementation of measures to limit the spread of the SARS-CoV-2 virus at the start of the COVID-19 pandemic resulted in a rapid decrease in all other respiratory pathogens. As COVID-19 containment measures were relaxed, the first non-COVID respiratory viruses to return to prepandemic levels were members of the rhinovirus/enterovirus, followed by the rapid return of seasonal coronaviruses, parainfluenza, and respiratory syncytial virus after the complete removal of COVID-19 precautions at the state level, including an end to mask mandates. Inasmuch as COVID-19 has dominated the landscape of respiratory infections since early 2020, it is important for clinicians to recognize the return of non-COVID respiratory pathogens may be rapid and significant when COVID-19 containment measures are removed.

## New Data Letter

Implementation of measures to limit SARS-CoV-2 transmission during the COVID-19 pandemic coincided with a marked decrease in infections caused by other respiratory viruses.(1) The decrease was due, in part, to masking, social distancing, closure of schools and businesses, and other efforts limiting disease spread by respiratory droplets.(2)

Early in the public health response to COVID-19 in Houston, the Houston Livestock Show and Rodeo, was cancelled on March 11^th^ 2020 and a citywide stay at home order was implemented on March 25^th^ 2020 (https://abc13.com/houston-rodeo-coronavirus-update-texas-cancellation-livestock-show-and/6003475/, https://www.houstontx.gov/mayor/press/2020/stay-home-work-safe-order.html, last accessed May 25, 2021). Subsequently, rates of influenza virus, respiratory syncytial virus (RSV), rhinovirus/enterovirus, and seasonal coronavirus infections, as diagnosed by the respiratory pathogen panel in the Houston Methodist Hospital centralized microbiology laboratory, declined rapidly (Figure 1A) (https://flu.houstonmethodist.org, last accessed: May 25, 2021).

**Figure 1.**
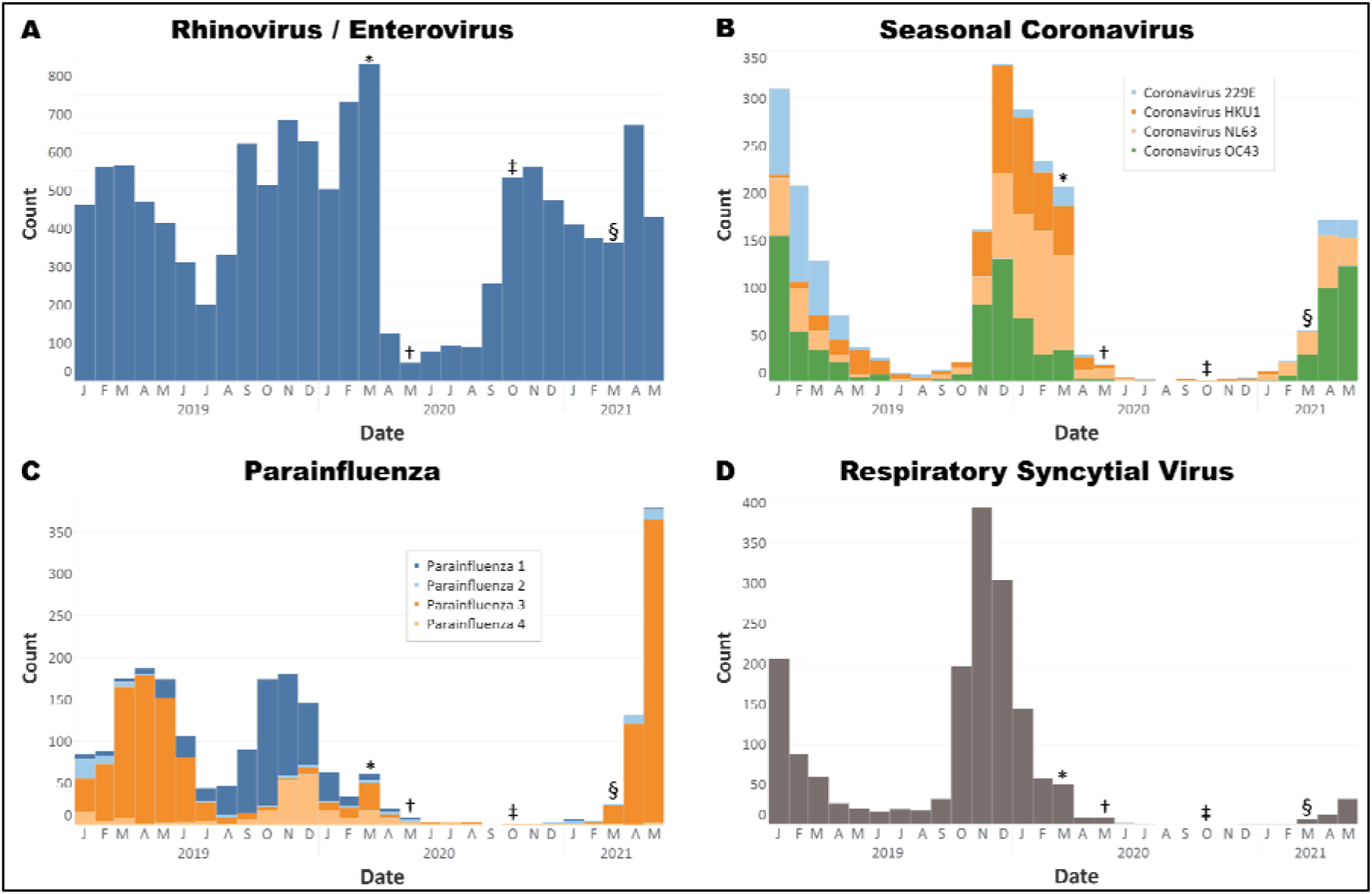
Histograms of non-COVID-19 positive respiratory virus tests from Jan 1, 2019 – May 25, 2021. A. Rhinovirus/enterovirus was the first virus to rebound to prepandemic levels. B. Coronaviruses 229E, HKU1, NL63, OC43. OC43 have been prevalent since March 2021. C. Parainfluenza viruses 1-4. Parainfluenza 3 has been prevalent since March 2021. D. Respiratory syncytial virus have also recently increased in prevalence. Symbols indicate the implementation and relaxation of COVID-19 precautions. * March 2020, start of pandemic lockdown measures. † May 2020, phase one reopening of Texas. ‡ October 2020, additional reopening measures. § March 2021, removal of all COVID restrictions, including elimination of mask mandates. All tests were performed on the Biofire Respiratory Pathogen Panel.

As COVID-19 measures were gradually relaxed starting in May 2020, very low levels of non-SARS-CoV-2 respiratory pathogens were detected even as cases of COVID-19 began to rise (https://gov.texas.gov/uploads/files/press/EO-GA-23_phase_two_expanding_opening_COVID-19.pdf, last accessed May 25, 2021).(3) Levels of non-COVID respiratory infections remained exceedingly low through the summer (10 – 28 rhinovirus/enterovirus cases per week (cpw), 0 – 4 cpw of influenza A and influenza B, 0 – 1 cpw of RSV). In September 2020 rhinovirus/enterovirus cases began increasing to pre-pandemic levels as schools reopened and many remaining measures were relaxed in October 2020 (97 – 156 rhinovirus/enterovirus cases per week, October through December 2020) (Figure 1A) (https://gov.texas.gov/uploads/files/press/EO-GA-32_continued_response_to_COVID-19_IMAGE_10-07-2020.pdf, last accessed May 25, 2021).

The first week of March 2021, the Texas governor announced remaining measures were being eliminated and face masks could no longer be mandated by state or local government (https://open.texas.gov/uploads/files/organization/opentexas/EO-GA-34-opening-Texas-response-to-COVID-disaster-IMAGE-03-02-2021.pdf, last accessed May 25, 2021). This same month, we began to observe a marked increase in rhinovirus/enterovirus, parainfluenza viruses, and seasonal coronavirus infections (Figure 1B-D). In Houston, seasonal coronaviruses typically peak during the winter months with very low levels observed during the summer. We are now observing a month-over-month increase in parainfluenza and seasonal coronaviruses which would be considered “out of season” compared to their typical seasonality. Out-of-season increases in RSV have been reported elsewhere when COVID-19 measures are relaxed (https://www.abc.net.au/news/2021-02-24/rsv-cases-surging-in-south-east-queensland/13186788, last accessed May 25, 2021). We have also seen a recent small increase in RSV cases in May 2021 (0 – 1 cpw between June 1 2020 and March 1 2020, now increased to 9 – 16 cpw in May 2021), although our patient population is primarily comprised of adults. Rhinovirus/enterovirus cases have also increased from 82 – 106 cpw in January and February 2021 to 143 to 156 cpw from the end of March through April 2021.

Inasmuch as the incremental relaxation of COVID-19 prevention measures affected respiratory infection rates over time, the recent discontinuation of mask mandates in March 2021 has coincided with a rapid increase in a variety of non-COVID respiratory pathogens. These observations are important for clinicians to consider as they evaluate patients with respiratory infections in the coming months. They also underscore the high effectiveness of non-pharmacologic preventative measures such as masking and social distancing in preventing the spread of respiratory pathogens spread by droplets.

## Data Availability

Data for influenza, RSV, and rhinovirus/enterovirus is available on flu.houstonmethodist.org. Data for non-COVID coronaviruses is available from authors upon request.

https://flu.houstonmethodist.org

